# A Normal Forced Vital Capacity Does Not Reliably or Equitably Exclude Restriction

**DOI:** 10.1101/2024.10.22.24315945

**Authors:** Alexander T. Moffett, Aparna Balasubramanian, Meredith C. McCormack, Jaya Aysola, Scott D. Halpern, Gary E. Weissman

**Affiliations:** Division of Pulmonary, Allergy, and Critical Care Medicine, Department of Medicine, University of Pennsylvania, Philadelphia, PA, USA; Palliative and Advanced Illness Research (PAIR) Center, University of Pennsylvania, Philadelphia, PA, USA; Leonard Davis Institute of Health Economics, University of Pennsylvania, Philadelphia, PA, USA; Division of Pulmonary and Critical Care Medicine, Johns Hopkins University, Baltimore, MD, USA; Penn Medicine Center for Health Equity Advancement, Office of the Chief Medical Officer, University of Pennsylvania Health System, Philadelphia, PA, USA; Division of General Internal Medicine, Department of Medicine, University of Pennsylvania, Philadelphia, PA, USA; Department of Biostatistics, Epidemiology, and Informatics, University of Pennsylvania, Philadelphia, PA, USA; Department of Medical Ethics and Health Policy, University of Pennsylvania, Philadelphia, PA, USA

## Abstract

**Background:** European Respiratory Society and American Thoracic Society (ERS/ATS) guidelines for pulmonary function test (PFT) interpretation recommend the use of a normal forced vital capacity (FVC) to exclude restriction. However, this recommendation is based upon a single study from 1999, which was limited to White patients, and used race-specific reference equations that are no longer recommended by ERS/ATS. We sought to reassess the support for this recommendation by calculating the negative predictive value (NPV) of a normal FVC in a diverse, multicenter cohort using race-neutral reference equations.

**Methods:** We interpreted PFTs performed between 2000 and 2023 in two academic medical systems and in a national electronic health record (EHR) database. We calculated the NPV of a normal FVC to exclude restriction overall and among pre-specified racial and ethnic groups.

**Results:** We included PFTs from 85 990 patients. The prevalence of restriction was 35.1%. The overall NPV of a normal FVC to exclude restriction was 80.5% (95% CI 80.1% to 80.8%), compared to an NPV of 97.6% cited in support of ERS/ATS guidelines. The NPV ranged from 65.2% (95% CI 64.4% to 66.0%) among non-Hispanic Black patients to 85.9% (95% CI 85.6% to 86.3%) among non-Hispanic White patients. This difference was largely attributable to lower FVC z-scores among non-Hispanic Black patients.

**Conclusions:** The NPV of a normal FVC is lower than has been previously reported and varies by race and ethnicity. The approach to PFT interpretation recommended by ERS/ATS guidelines results in the under-recognition of restriction, particularly among non-Hispanic Black patients.

## Introduction

European Respiratory Society and American Thoracic Society (ERS/ATS) guidelines for pulmonary function test (PFT) interpretation recommend the use of a normal forced vital capacity (FVC) to exclude restriction.^1^ According to these guidelines, static lung volume measurements are needed to determine the presence or absence of restriction only if the FVC is abnormal. If the FVC is normal, restriction can be excluded from spirometry alone. In support of this recommendation, ERS/ATS guidelines cite a single study from 1999 which reported that the negative predictive value (NPV) of a normal FVC to exclude restriction is 97.6%.^2^ This study, however, involved fewer than two thousand PFTs performed at a single pulmonary diagnostic lab, included only White patients, and employed race-specific reference equations that are no longer recommended by ERS/ATS.^3–5^ While subsequent studies have provided similar NPV estimates, these studies have also been limited to small cohorts of patients tested at a single site.^6,7^ Moreover, no study has assessed the effect of adopting the recommended race-neutral Global Lung Function Initiative (GLI) Global reference equations,^8^ despite the significant impact these equations have on the FVC lower limit of normal (LLN).^9–11^

We sought to evaluate the validity and equity of current ERS/ATS guidelines by reassessing the NPV of a normal FVC in a large, diverse, multi-center cohort, using race-neutral reference equations, and comparing the NPV among different racial and ethnic groups.

## Methods

### Study Population

We included PFTs with both static and dynamic lung volume measurements performed between 2000 and 2023 at four pulmonary diagnostic labs within Johns Hopkins Medicine and three pulmonary diagnostic labs within Penn Medicine. We also included PFTs with both static and dynamic lung volumes, performed between 2007 and 2023 and reported in the Optum Labs Data Warehouse (OLDW), a database of electronic health record (EHR) data collected from across the United States.^12,13^ We excluded PFTs from patients younger than 18 years of age and older than 80 years of age, with the latter limitation imposed by the use of GLI 2019 reference equations for static lung volumes.^14^ For patients with multiple PFTs we limited our analysis to the first PFT performed for each patient. For the Johns Hopkins Medicine and Penn Medicine PFT data, patient race and ethnicity were as documented on the PFT report. Minimal granularity was seen in the Penn Medicine PFT data—with patients classified as Asian, Hispanic, non-Hispanic Black, non-Hispanic White, and Other— and this classification was applied to the Johns Hopkins and OLDW data. This set of racial and ethnic categories reflects the historical use of race-specific reference equations in which normal pulmonary function was thought to differ across Hispanic, non-Hispanic Black, and non-Hispanic white patients.^15^

### Pulmonary Function Tests

Pulmonary function tests at Johns Hopkins Medicine and Penn Medicine were performed in accordance with ATS recommendations.^16–19^ Static lung volumes were measured with either helium dilution or plethysmography at the pulmonary diagnostic labs within Johns Hopkins Medicine and with plethysmography at the labs within Penn Medicine. Technical data regarding the performance of the PFTs included in the OLDW were not available.

PFTs were interpreted using 2021 ERS/ATS guidelines.^1^ FVC z-scores were calculated using race-neutral GLI Global equations, while TLC z-scores were calculated using GLI 2019 equations for static lung volumes.^8,14^ A parameter value was normal if its z-score was greater than *−*1.645. Restriction was present if the TLC was abnormal.^1^ In a sensitivity analysis we considered the effect of interpreting PFTs using other reference equations.^3–5,15,20,21^

### Outcomes

The primary outcome was the NPV associated with the use of a normal FVC to exclude restriction. We estimated both the overall NPV and the NPV by patient race and ethnicity. To test if normal FVC values close to the LLN were associated with a lower NPV than FVC values farther from the LLN, we binned patients by FVC z-score in intervals of 0.2 and compared the NPV across these binned groups.

To assess whether differences in FVC z-score by race and ethnicity might account for racial and ethnic differences in NPV, we fit a logistic regression model to estimate the association between these demographic categories and the odds of restriction in patients with a normal FVC. We then compared this unadjusted model to a model adjusted by the FVC z-score.

To assess the effect of replacing the FVC z-score cutoff of *−*1.645 recommended by ERS/ATS guidelines, we calculated the NPV associated with other potential FVC z-score cutoffs ranging from *−*3 to 0.5. We compared the NPVs associated with these alternate cutoffs by race and ethnicity and identified the cutoff values that would be needed to yield both an overall NPV of 97.6% and an NPV of at least 97.6% in each racial and ethnic group. We further compared the effect that these cutoffs would have on the number of patients for whom static lung volume measurements were recommended.

### Statistical Analysis

All analyses were performed using R version 4.4.1.^22^ Confidence intervals for proportions were calculated using the Clopper-Pearson exact method.^23^ All statistical tests were two sided and a *P* value *<* 0.05 was interpreted as statistically significant.

This study was performed in accordance with the Strengthening the Reporting of Observational Studies in Epidemiology (STROBE) reporting guidelines (**Table S1**).^24^ The study was approved by the Johns Hopkins University and University of Pennsylvania Institutional Review Boards.

## Results

We interpreted PFTs from 85 990 patients (**Table 1**). Most PFTs were from female (50 168 [58.3%]) and from non-Hispanic White (52 345 [60.9%]) or non-Hispanic Black (24 272 [28.2%]) patients. The mean age was 56.8 years (standard deviation [SD] 14.5). The mean FVC z-score was *−*0.9 (SD 1.4). The mean TLC z-score was *−*1.2 (SD 1.6). Restriction was present in 30 254 (35.2%) patients. Significant variation in pulmonary function was seen across different racial and ethnic groups, with FVC and TLC z-scores significantly lower, and the prevalence of restriction significantly higher, among non-Hispanic Black patients (**Table S2-S4**). Significant variation was also seen across different pulmonary diagnostic labs, with the prevalence of restriction ranging from 17.7% to 39.7% in PFTs performed at Penn Medicine (**Table S5**).

**Table 1:** Patient Characteristics.

|  | Dataset |  |  |  |
| --- | --- | --- | --- | --- |
|  | Johns Hopkins<br>Medicine<br>(*n* = 38 989) | Penn<br>Medicine<br>(*n* = 41 926) | OLDW<br>(*n* = 5 075) | All<br>(*n* = 85 990) |
| **Age, years** | 57.2 (14.3) | 55.8 (14.6) | 62.2 (13.2) | 56.8 (14.5) |
| **Sex** |  |  |  |  |
| Male | 16 339 (41.9) | 17 374 (41.4) | 2 109 (41.6) | 35 822 (41.7) |
| Female | 22 650 (58.1) | 24 552 (58.6) | 2 966 (58.4) | 50 168 (58.3) |
| **Race and Ethnicity** |  |  |  |  |
| Asian | 458 (1.2) | 516 (1.2) | 95 (1.9) | 1 069 (1.2) |
| Hispanic | 37 (0.1) | 570 (1.4) | 187 (3.7) | 794 (0.9) |
| Non-Hispanic Black | 10 191 (26.1) | 13 662 (32.6) | 419 (8.3) | 24 272 (28.2) |
| Non-Hispanic White | 22 540 (57.8) | 25 759 (61.4) | 4 046 (79.7) | 52 345 (60.9) |
| Other | 5 763 (14.8) | 1 419 (3.4) | 328 (6.5) | 7 510 (8.7) |
| **Dynamic Lung Volumes, z-score** |  |  |  |  |
| FEV<sub>1</sub> | −1.1 (1.5) | −1.4 (1.4) | −0.6 (1.4) | −1.2 (1.5) |
| FVC | −0.8 (1.4) | −1.1 (1.3) | −0.3 (1.3) | −0.9 (1.4) |
| FEV<sub>1</sub>/FVC | −0.6 (1.3) | −0.6 (1.5) | −0.7 (1.6) | −0.6 (1.4) |
| **Static Lung Volumes, z-score** |  |  |  |  |
| TLC | −1.5 (1.6) | −0.9 (1.5) | −0.6 (1.4) | −1.2 (1.6) |
| **Interpretation** |  |  |  |  |
| Normal | 16192 (41.5) | 18502 (44.1) | 3251 (64.1) | 37945 (44.1) |
| Non-Specific | 924 (2.4) | 2776 (6.6) | 19 (0.4) | 3719 (4.3) |
| Obstructive | 5305 (13.6) | 7949 (19.0) | 818 (16.1) | 14072 (16.4) |
| Restrictive | 14227 (36.5) | 11520 (27.5) | 861 (17.0) | 26608 (30.9) |
| Mixed | 2341 (6.0) | 1179 (2.8) | 126 (2.5) | 3646 (4.2) |
| **Severity** |  |  |  |  |
| Normal | 25583 (65.6) | 25168 (60.0) | 3859 (76.0) | 54610 (63.5) |
| Mild | 6674 (17.1) | 8153 (19.4) | 630 (12.4) | 15457 (18.0) |
| Moderate | 5842 (15.0) | 7202 (17.2) | 527 (10.4) | 13571 (15.8) |
| Severe | 890 (2.3) | 1403 (3.3) | 59 (1.2) | 2352 (2.7) |

Values are means (SD) for continuous variables and counts (percentage) for categorical variables. Dynamic lung volume z-scores are calculated using GLI Global reference equations. Static lung volume z-scores are calculated using GLI 2019 reference equations. Abbreviations: FEV<sub>1</sub> = forced expiratory volume in 1 second; FVC = forced vital capacity; GLI = Global Lung Function Initiative; LLN = lower limit of normal; SD = standard deviation; TLC = total lung capacity.

The overall NPV associated with the use of a normal FVC to exclude restriction was 80.5% (95% confidence interval [CI] 80.1% to 80.8%). This was substantially lower than the NPV of 97.6% cited by ERS/ATS in support of current guidelines and lower than the NPV reported in subsequent studies (**Table 2**).^2^ Significant variation was seen in the NPV of a normal FVC across the different datasets, with the NPV ranging from 72.9% (95% CI 72.4% to 73.5%) in the Johns Hopkins Medicine dataset, to 86.7% (95% CI 86.3% to 87.1%) in the Penn Medicine dataset, and 89.0% (88.1% to 90.0%) in the OLDW dataset (**Table S6**). The NPV varied significantly across different pulmonary diagnostic labs at Penn Medicine, ranging from 80.0% (78.8% to 81.3%) to 95.0% (94.3% to 95.6%) (**Table S7**).

**Table 2:**
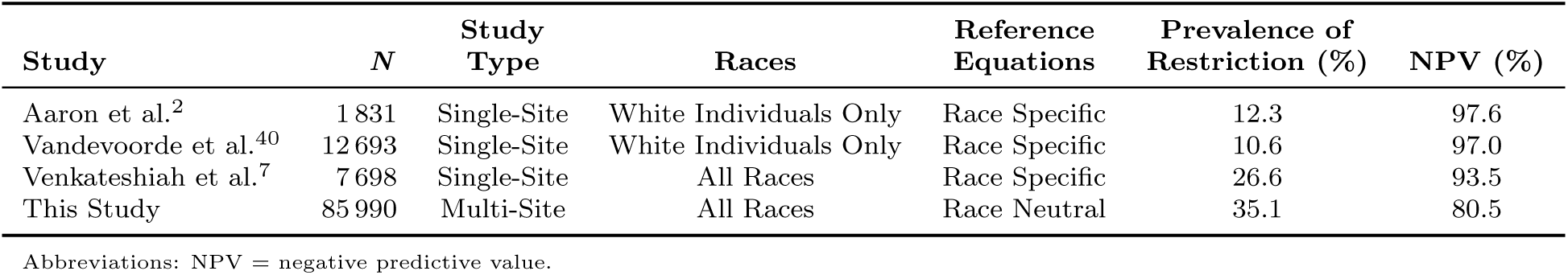
The NPV of a Normal FVC to Exclude a Restriction in This and Other Studies.

Though FVC and TLC z-scores were correlated, a given FVC z-score was associated with a wide range of TLC z-scores (**Figure 1**). A substantial fraction of normal FVC z-scores near the LLN were thus associated with abnormal TLC z-scores (**Figure 2**). In the Penn Medicine and Johns Hopkins Medicine datasets the NPV ranged from 44.9% (95% CI 44.3% to 46.4%) for patients with a z-score in the interval [−1.645, −1.445) to 99.5% (95% CI 98.9% to 99.9%) for patients with a z-score in the interval [0.955, 1.155).

**Figure 1.**
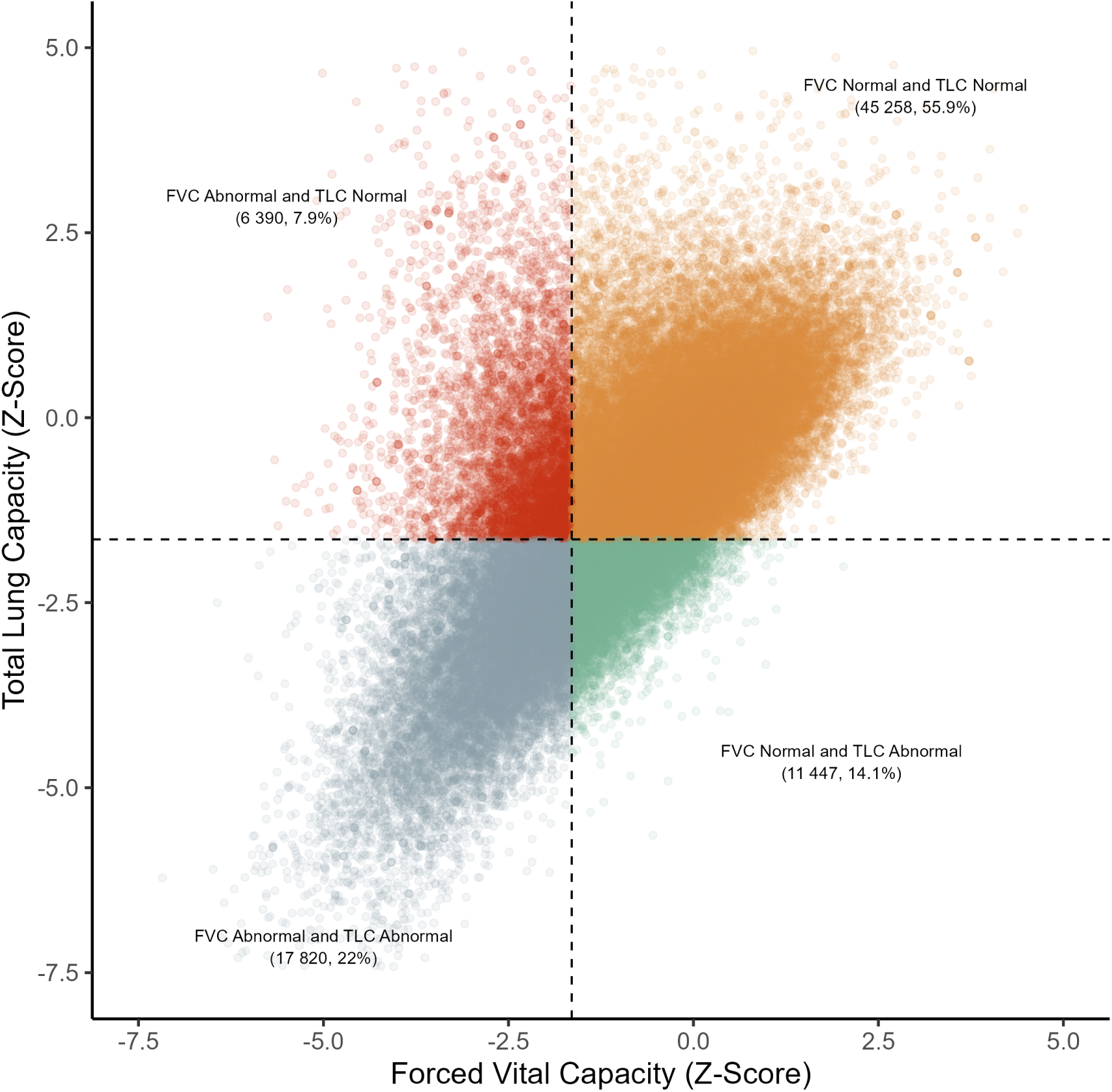
Association of FVC z-scores and TLC z-scores. FVC and TLC values are drawn from 80 915 PFTs performed at Johns Hopkins Medicine and Penn Medicine. FVC z-scores were calculated using GLI Global reference equations. TLC z-scores were calculated using GLI 2019 equations. Dashed lines represent the lower limit of normal z-score cutoff of −1.645. FVC = forced vital capacity; GLI = Global Lung Function Initiative; PFT = pulmonary function test; TLC = total lung capacity.

**Figure 2.**
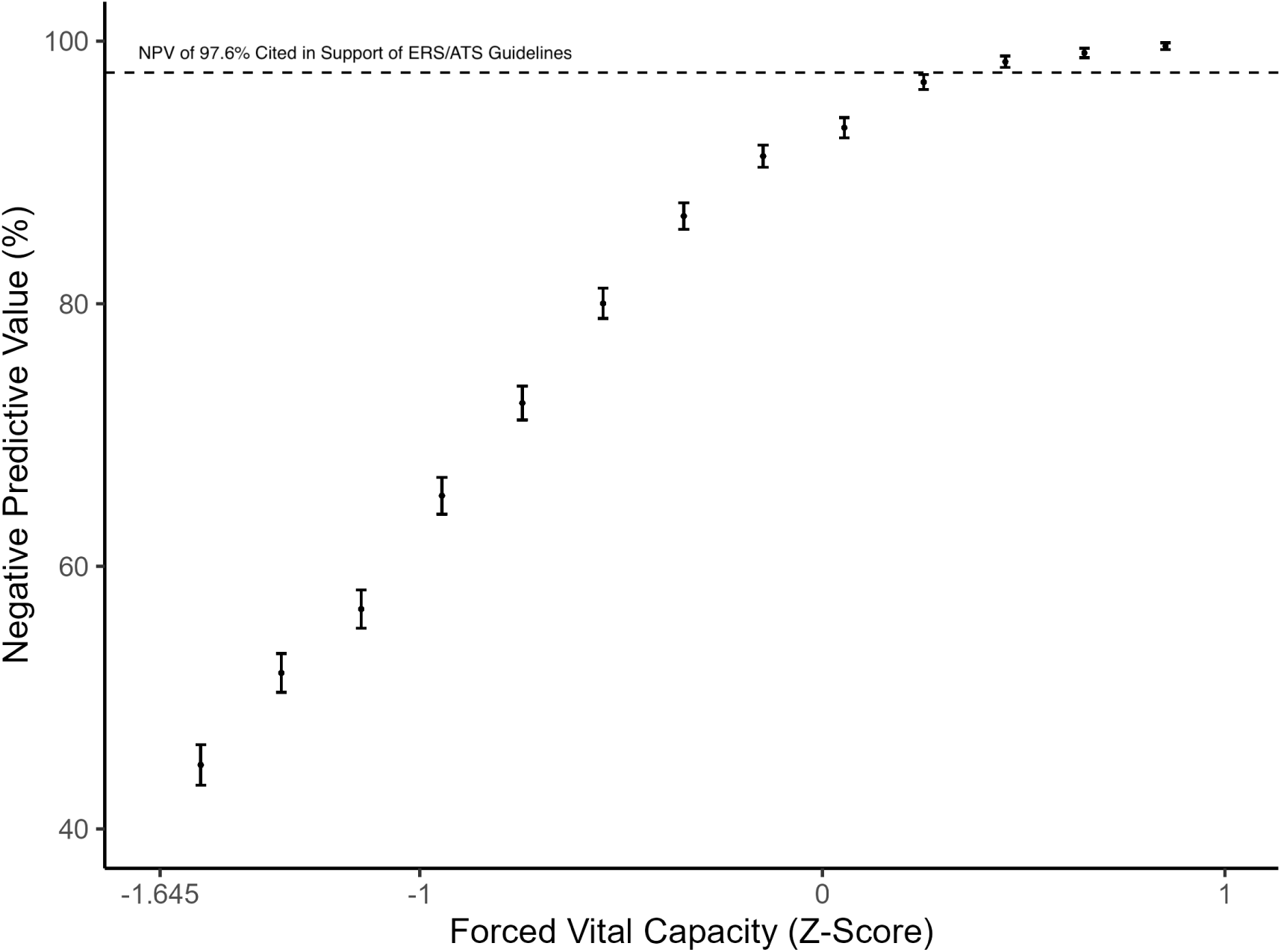
Association between FVC z-scores and the NPV of a normal FVC to exclude restriction. FVC and TLC z-scores were drawn from 80 915 PFTs performed at Johns Hopkins Medicine and Penn Medicine. FVC z-scores were calculated using GLI Global reference equations while TLC z-scores were calculated using GLI 2019 equations. FVC z-scores were binned in intervals of 0.2 and the NPV of a normal FVC to exclude a restriction was calculated for each bin. Intervals represent 95% confidence intervals. The dotted line corresponds to the NPV of 97.6% cited by ERS/ATS in support of current guidelines. ATS = American Thoracic Society; ERS = European Respiratory Society; FVC = forced vital capacity; GLI = Global Lung Function Initiative; NPV = negative predictive value; PFT = pulmonary function test; TLC = total lung capacity.

The NPV varied substantially by race and ethnicity, ranging from 65.2% (95% CI 64.4% to 66.0%) among non-Hispanic Black patients to 85.9% (95% CI 85.6% to 86.3%) among non-Hispanic White patients (**Figure 3**). In the Johns Hopkins, Penn, and OLDW datasets the NPV of non-Hispanic Black patients was 52.1% (95% CI 50.8% to 53.5%), 75.8% (95% CI 74.8% to 76.8%), and 62.2% (95% CI 56.1% to 68.0%), respectively (**Table S8**).

**Figure 3.**
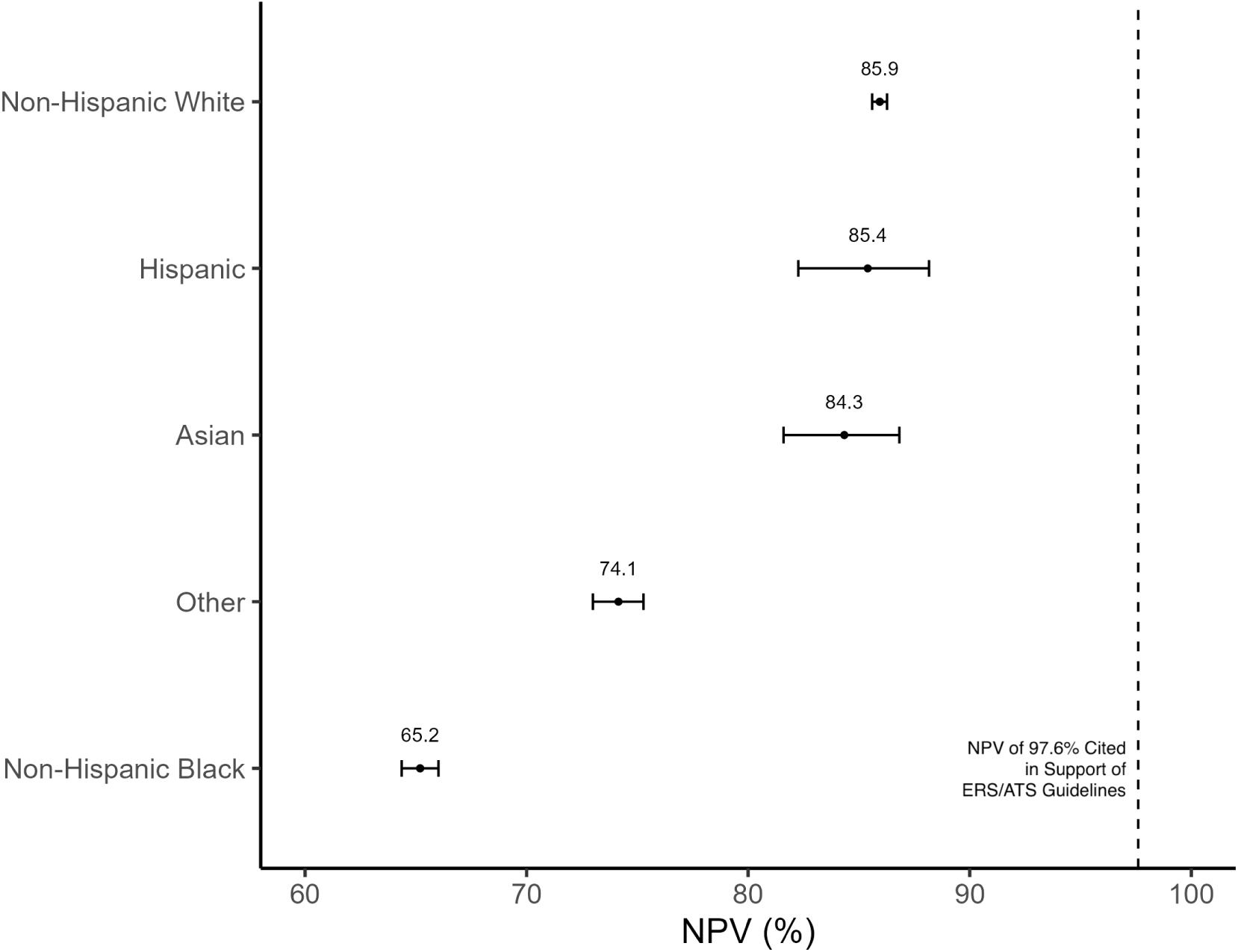
Variation in the NPV of a normal FVC to exclude restriction by race and ethnicity. FVC and TLC z-scores are drawn from 85 990 PFTs from Johns Hopkins Medicine, Penn Medicine, and the Optum Labs Data Warehouse. FVC z-scores were calculated using GLI Global reference equations while TLC z-scores were calculated using GLI 2019 equations. Intervals represent 95% confidence intervals. The dashed line corresponds to the NPV of 97.6% cited by ERS/ATS in support of current guidelines. ATS = American Thoracic Society; ERS = European Respiratory Society; FVC = forced vital capacity; NPV = negative predictive value; PFT = pulmonary function test; TLC = total lung capacity.

The lower NPV among non-Hispanic Black patients was largely a consequence of lower FVC z-scores among these patients. In the unadjusted logistic regression model, a normal FVC among non-Hispanic Black patients in the Johns Hopkins and Penn datasets was associated with a 3.1 (95% CI 3.0 to 3.3) times increased odds of having an abnormal TLC relative to non-Hispanic White patients (**Table 3**). After adjusting for FVC z-score, this odds ratio decreased to 1.6 (95% CI 1.5 to 1.6). Further adjusting for age and sex had no additional effect on this estimate.

**Table 3:**
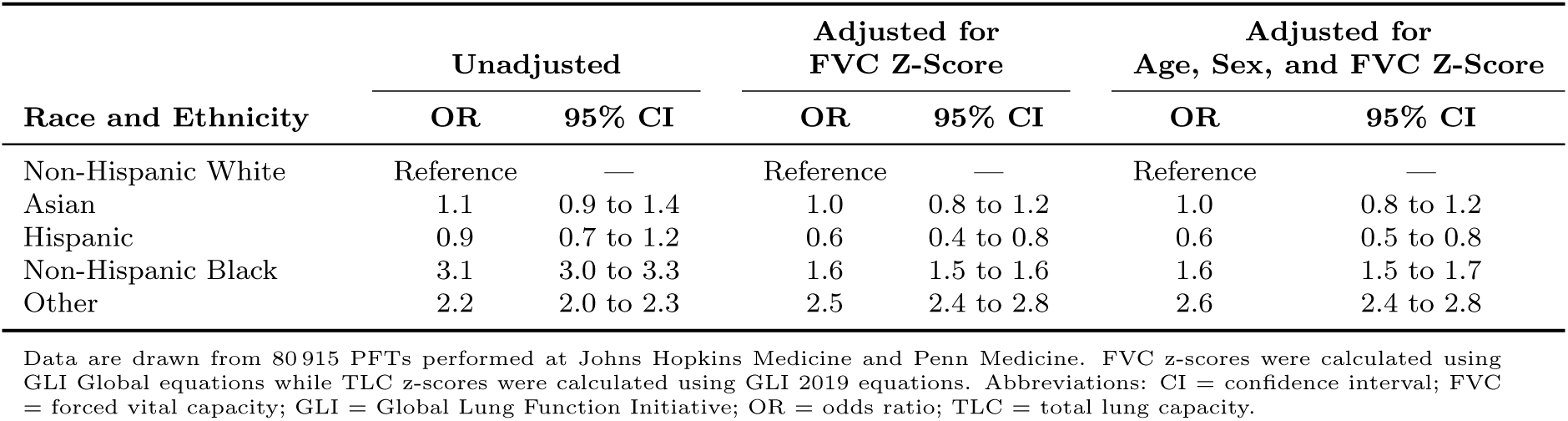
Differences by Race and Ethnicity in the Probability that a Normal FVC is Associated with an Abnormal TLC.

The choice of reference equations had a significant effect on the NPV in the Johns Hopkins Medicine and Penn Medicine datasets (**Table S9**). Applying the same reference equations used to arrive at an NPV of 97.6% to the non-Hispanic White patients in these datasets increased the NPV from 85.4% to 92.2% (95% CI 92.0% to 92.5%).^2–5^ The NPV varied from 84.7% (95% CI 84.3% to 85.1%) to 95.8% (95% CI 95.5% to 96.0%) across other reference equations.^3,8,15,20^ When applied to patients of all races in the Penn Medicine and Johns Hopkins Medicine datasets, the NPV of the race-neutral GLI Global equations was 79.8% (95% CI 79.5% to 80.1%), while the NPV of the race-specific GLI 2012 equations was 78.8% (95% CI 78.4% to 79.1%) (**Table S10**). Among non-Hispanic Black patients, the race-neutral equations had an NPV of 65.3% (95% CI 64.4% to 66.1%) while the race-specific equations had an NPV of 58.1 (95% CI 57.3% to 58.8%).

To ensure an overall NPV of 97.6% in the Johns Hopkins Medicine and Penn Medicine datasets, the FVC z-score cutoff of *−*1.645 recommended by current ERS/ATS guidelines would need to be increased to *−*0.083 (**Figure 4**), while to ensure an NPV of at least 97.6% among each racial and ethnic group, the z-score cutoff would need to be further increased to 0.112. Applying the *−*0.083 cutoff to these datasets would increase the number of patients for whom static lung volume measurements were recommended from 24 210 (29.9%) to 58 727 (72.6%), while applying the 0.112 cutoff would further increase the number of recommended static lung volume measurements to 62 605 (77.4%) (**Figure 5**). Among non-Hispanic Black patients, applying the 0.112 cutoff would increase the number of patients for whom static lung volume measurements were recommended from 11 375 (47.7%) to 22 560 (94.6%).

**Figure 4.**
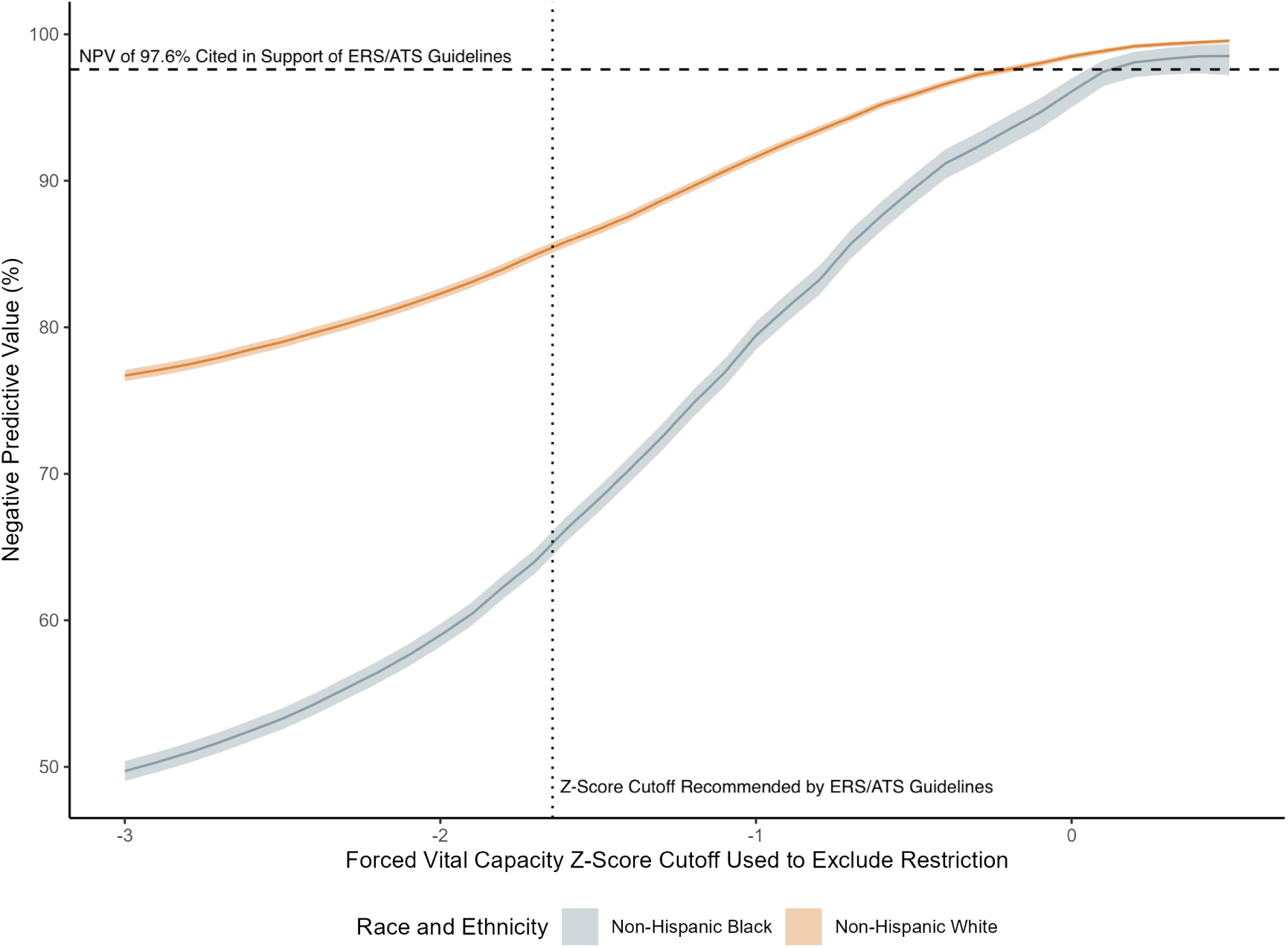
Association between the choice of FVC z-score cutoff and the NPV by race and ethnicity. FVC and TLC z-scores are drawn from 80 915 PFTs performed at Johns Hopkins Medicine and Penn Medicine. The x-axis represents different potential z-score cutoffs with which to exclude the presence of restriction while the y-axis represents the NPV associated with each cutoff, by patient race and ethnicity. The bands represent 95% confidence intervals. The dashed line corresponds to the NPV of 97.6% cited in support of ERS/ATS guidelines while the dotted line corresponds to the FVC z-score cutoff of *−*1.645 recommended by these guidelines. ATS = American Thoracic Society; ERS = European Respiratory Society; FVC = forced vital capacity; NPV = negative predictive value; PFT = pulmonary function test; TLC = total lung capacity.

**Figure 5.**
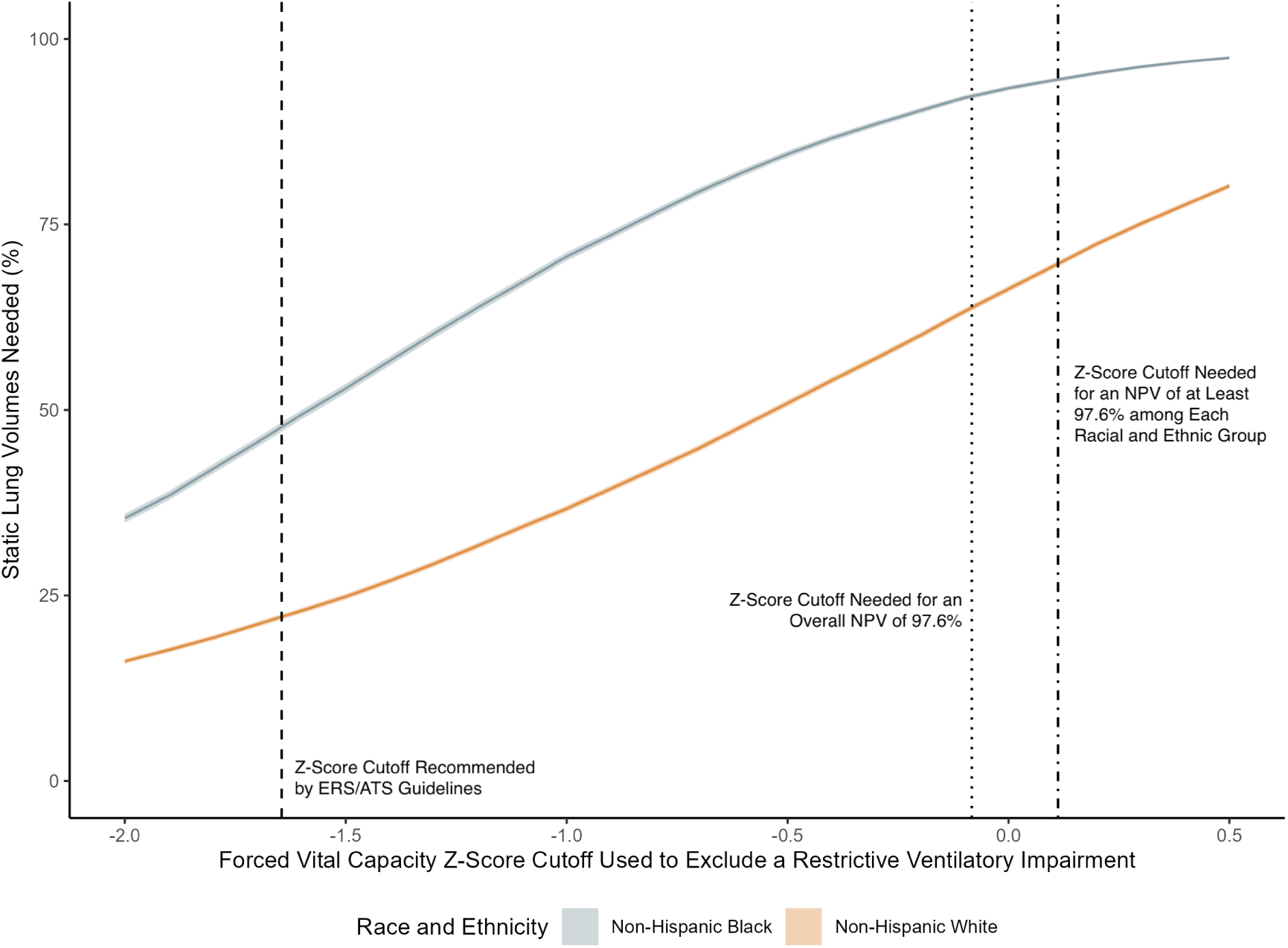
Association between the choice of FVC z-score cutoff and the percentage of PFTs for which static lung volume measurements are needed. FVC and TLC z-scores are drawn from 80 915 PFTs performed at Johns Hopkins Medicine and Penn Medicine. The x-axis represents different potential z-score cutoffs with which to exclude the presence of restriction while the y-axis represents the percentage of tests for which static lung volume measurements are recommended on the basis of these cutoffs, by race and ethnicity. The bands represent 95% confidence intervals. The dashed line corresponds to the FVC z-score cutoff of *−*1.645 recommended by ERS/ATS guidelines. The dotted line corresponds to the cutoff of *−*0.083 needed for an overall NPV of 97.6% in the Johns Hopkins and Penn Medicine PFT data, while the dashed and dotted line corresponds to the cutoff of 0.112 needed for an NPV of at least 97.6% in each racial and ethnic group. ATS = American Thoracic Society; ERS = European Respiratory Society; FVC = forced vital capacity; NPV = negative predictive value; PFT = pulmonary function test; TLC = total lung capacity.

## Discussion

Applying race-neutral reference equations to PFTs drawn from two health systems and a national EHR database, we estimated the NPV of a normal FVC to exclude restriction to be 80.5%, which is considerably less than the estimate of 97.6% used to justify current ERS/ATS guidelines. We found the NPV to vary by patient race and ethnicity, with a significantly lower NPV among non-Hispanic Black patients. While ERS/ATS guidelines for PFT interpretation recommend the use of a normal FVC to exclude restriction, our findings suggest that this practice may lead to the missed or delayed identification of restriction in a significant percentage of tests, particularly among non-Hispanic Black patients.

Selecting an FVC z-score cutoff with which to exclude restriction involves weighing the cost of failing to identify restriction against the benefit of limiting the need for static lung volume measurements. While this study has not identified the optimal cutoff with which to exclude restriction—as this would depend upon an assessment of patient and physician values and preferences—it has demonstrated that current ERS/ATS guidelines are based on an inaccurate account of the probability that a normal FVC will falsely exclude restriction. To produce an overall NPV of 97.6% in our cohort, the z-score cutoff used to exclude the presence of a restriction would need to be increased from *−*1.645 to *−*0.083, a change that would more than double the number of patients for whom static lung volume measurements are recommended.

The overall NPV in our study was substantially lower than that reported in other studies.^2,6,7^ This is likely due both to differences in the reference equations used in our study and to differences in the distribution of FVC z-scores in our cohort. We found the NPV to depend upon the choice of reference equations and that the specific equations used to produce an NPV estimate of 97.6% yielded a higher estimate among non-Hispanic White patients than the GLI global reference equations recommended by ERS/ATS and employed in this study. If other studies had used the GLI Global equations, their NPV estimates would have been lower. At the same time, we found that the probability that a normal FVC will accurately predict the presence of a normal TLC depends on the distance of the FVC z-score from the LLN cutoff. The NPV of an FVC just above the LLN was less than 50%, while the NPV approached 100% with FVC z-scores greater than 0. Cohorts in which the mean FVC z-score is closer to *−*1.645 will have a lower overall NPV than cohorts in which the FVC z-score is closer to 0.

The lower overall NPV reported in our study represents an example of dataset shift, the phenomenon in which a clinical prediction model developed in one clinical setting sees a decline in performance when it is applied in a different setting.^25–27^ In this case, a simple prediction model—that a normal FVC predicts the presence of a normal TLC—sees a decline in performance when applied to a different clinical population with different reference equations. This study highlights the importance both of assessing such performance in large, diverse cohorts, and of reassessing such performance in response to the adoption of novel reference equations.

In addition to a lower overall NPV, we also found significant variation in the NPV of a normal FVC across different racial and ethnic groups. The odds that restriction was present in a patient with a normal FVC were more than three times higher in non-Hispanic Black patients than in non-Hispanic White patients. This unequal performance is largely a result of differences in the distribution of FVC z-scores across those racial and ethnic groups; as non-Hispanic Black patients had lower FVC z-scores than patients of other races and ethnicities, their PFTs were more likely to be misinterpreted. In this way, the structural inequities responsible for the observed differences in FVC z-score by race—differences in exposure to environmental^28,29^ and socioeconomic^30–32^ risk factors along with differences in access to care^33,34^—are reinforced by the use of a normal FVC to exclude restriction. Restriction is more likely to be missed in non-Hispanic Black patients, leading to suboptimal care for these patients and thus to even greater disparities in respiratory health.^35–37^

It should be noted that the unequal performance across different racial and ethnic groups is present despite the use of race-neutral reference equations. Indeed, when compared to the race-specific GLI 2012 equations, the use of race-neutral GLI Global equations was associated with an improvement in both the overall NPV of a normal FVC, and the NPV of a normal FVC among non-Hispanic Black patients. The replacement of race-specific with race-neutral reference equations illuminates the significant differences, at the time of testing, in the pulmonary function of non-Hispanic Black patients when compared to that of patients of other races and ethnicities. Such differences must be considered, when developing an approach to PFT interpretation, to reduce the likelihood of bias.

This study has several strengths. First, we interpreted a larger number of PFTs than past studies, drawn from a diverse patient population. While previous studies have been limited to the assessment of PFTs performed at a single pulmonary diagnostic lab, our data were drawn from two health systems and a national EHR database. Second, we provided the first estimate of the effect of race-neutral reference equations on the NPV of a normal FVC to exclude restriction and compared the use of these equations with those of other reference equations, demonstrating the effect of the choice of reference equations on the NPV of a normal FVC. Third, we provided the first assessment of variation in the NPV of a normal FVC across racial and ethnic groups and identified a novel and important source of inequity in the approach to PFT interpretation recommended by ERS/ATS.

This study also has limitations. First, there is the potential for ascertainment bias. Our cohort was necessarily limited to PFTs that included both static and dynamic lung volumes, yet the results have the greatest implications for the interpretation of PFTs that include only dynamic lung volume measurements, and the corresponding clinical management of such patients (e.g., whether to then obtain static measurements). However, this limitation also applies to all other studies that have estimated the NPV of a normal FVC, and so are unlikely to explain the much lower NPV we observe. Further, this limitation would not explain the dramatic variability we observe in the NPV across racial groups. Second, the racial and ethnic categories used in this study reflect the racial and ethnic categories documented in PFT reports, and we were unable to use the set of racial categories recommended by the National Institutes of Health, or to separate race from ethnicity. Moreover, the racial and ethnic data in the OLDW were imputed, rather than self-reported.^38,39^ Third, while the PFTs included in the OLDW were drawn from a national database, PFT data were included for only a subset of those patients who had likely undergone testing, with the selection effects responsible for such inclusion potentially limiting the external validity of our findings. Fourth, while FVC z-scores were calculated with race-neutral reference equations, the GLI 2019 reference equations for static lung volumes were derived from individuals of European ancestry.^14^ Reference equations developed from a more diverse population are not available for static lung volumes. Fifth, our study considered only the impact of ERS/ATS guidelines on PFT interpretation and did not further assess the downstream clinical consequences of these guidelines. While ERS/ATS guidelines state that static lung volume measurements are not needed to assess for restriction in patients with a normal FVC, it is unclear to what extent this recommendation is followed in practice.

In conclusion, applying race-neutral reference equations to a diverse multicenter cohort, we found that the NPV of a normal FVC to exclude restriction is substantially less than has been reported, with significant variation by patient race and ethnicity.

## Supporting information

Supplemental Online Content

## Data Availability

All data produced in the present study are available upon reasonable request to the authors.

## Notes

### Competing Interest Statement

The authors have declared no competing interest.

### Funding Statement

ATM reports funding from NHLBI F32 HL167456. GEW reports funding from NHLBI R03 HL171424.

### Author Declarations

The study was approved by the Johns Hopkins University and University of Pennsylvania Institutional Review Boards.

