## Supplemental Online Content for "A Normal Forced Vital Capacity Does Not Reliably or Equitably Exclude Restriction"


__Tables__

**Table S1.** STROBE Checklist.

**Table S2.** Patient Characteristics in the Johns Hopkins Medicine Dataset by Race and Ethnicity

**Table S3.** Patient Characteristics in the Penn Medicine Dataset by Race and Ethnicity

**Table S4**. Patient Characteristics in the OLDW Dataset by Race and Ethnicity

**Table S5.** Patient Characteristics by Pulmonary Diagnostic Lab

**Table S6.** Test Characteristics by Dataset

**Table S7.** Test Characteristics by Pulmonary Diagnostic Lab

**Table S8.** Test Characteristics by Race and Ethnicity

**Table S9.** NPV in Non-Hispanic White Patients with Different Reference Equations

**Table S10.** Test Characteristics with Race-Specific and Race-Neutral Reference Equations

__Figures__

**Figure S1** Flow Diagram for the Johns Hopkins Medicine Dataset

**Figure S2** Flow Diagram for the Penn Medicine Dataset

**Figure S3** Flow Diagram for the OLDW Dataset

__References__

**Supplementary Table S1**. STROBE Checklist

| | **Item** | **Recommendation** | **Location** |
|---|---|---|---|
| Title and Abstract | 1a | Indicate study design | Page 1 |
| | 1b | Informative and balanced summary | Pages 1–2 |
| | | *Introduction* | |
| Background and Rationale | 2 | Explain background and rationale | Page 3 |
| Objectives | 3 | State specific objectives | Page 3 |
| | | *Methods* | |
| Study Design | 4 | Present key elements | Page 5 |
| Setting | 5 | Describe setting, locations, and relevant dates | Page 4 |
| Participants | 6 | Give eligibility criteria and selection | Page 4 |
| Variables | 7 | Define outcomes, exposures, and predictors | Page 5 |
| Data Sources and Measurements | 8 | Give source and assessment of each variable | Pages 4–5 |
| Bias | 9 | Describe efforts to address bias | Pages 4–5 |
| Study Size | 10 | Explain how the study size was determined | N/A |
| Quantitative Variables | 11 | Explain handling of quantitative variables | Pages 4–5 |
| Statistical Methods | 12a | Describe all statistical methods | Page 6 |
| | 12b | Describe methods for subgroups and interactions | Page 4 |
| | 12c | Explain how missing data were addressed | Figures S1–S3 |
| | 12d | Describe sampling strategy | N/A |
| | 12e | Describe sensitivity analysis | Page 4, Table S9 |
| | | *Results* | |
| Participants | 13a | Report number of individuals at each stage of study | Figures S1–S3 |
| | 13b | Give reasons for non-participation at each stage | Figure S1–S3 |
| | 13c | Consider use of a flow diagram | Figures S1–S3 |
| Descriptive Data | 14a | Give characteristics of study participants | Tables 1, S2–S5 |
| | 14b | Indicate number of participants with missing data | Figures S1–S3 |
| Outcome Data | 15 | Report number of outcome events | Table 1 |
| Main Results | 16a | Give adjusted and unadjusted estimates | Table 3 |
| | 16b | Report category boundaries | N/A |
| | 16c | Translate relative risk into absolute risk | N/A |
| Other Analyses | 17 | Report other analyses performed | Page 5 |
| | | *Discussion* | |
| Key Results | 18 | Summarize key results | Page 10 |
| Limitations | 19 | Discuss limitations of the study | Pages 12–13 |
| Interpretation | 20 | Give an overall interpretation | Page 13 |
| Generalizability | 21 | Discuss external validity | Page 13 |
| | | *Other Information* | |
| Funding | 22 | Give source of funding | Title Page |

**Supplementary Table S2**. Patient Characteristics in the Johns Hopkins Medicine Dataset by Race and Ethnicity

| | **Asian** ($n = 458$) | **Hispanic** ($n = 37$) | **Non-Hispanic Black** ($n = 10\,191$) | **Non-Hispanic White** ($n = 22\,540$) | **Other** ($n = 5\,763$) |
|---|---|---|---|---|---|
| **Age, years** | 52.4 (15.9) | 49.7 (14.9) | 54.8 (13.5) | 58.9 (14.0) | 55.0 (15.5) |
| **Sex** | | | | | |
| Male | 179 (39.1) | 13 (35.1) | 3 569 (35.0) | 10 100 (44.8) | 2 478 (43.0) |
| Female | 279 (60.9) | 24 (64.9) | 6 622 (65.0) | 12 440 (55.2) | 3 285 (57.0) |
| **Dynamic Lung Volumes, z-score** | | | | | |
| $FEV_1$ | −1.1 (1.4) | −1.4 (1.4) | −1.7 (1.2) | −0.8 (1.5) | −0.9 (1.4) |
| FVC | −1.0 (1.4) | −1.3 (1.5) | −1.6 (1.2) | −0.5 (1.4) | −0.6 (1.4) |
| $FEV_1$/FVC | −0.5 (1.3) | −0.4 (0.9) | −0.6 (1.4) | −0.7 (1.3) | −0.5 (1.2) |
| **Static Lung Volumes, z-score** | | | | | |
| TLC | −1.6 (1.6) | −2.1 (2.0) | −2.3 (1.4) | −1.1 (1.5) | −1.5 (1.5) |
| **Interpretation** | | | | | |
| Normal | 198 (43.2) | 11 (29.7) | 2 172 (21.3) | 11 236 (49.8) | 2 575 (44.7) |
| Non-Specific | 10 (2.2) | 2 (5.4) | 312 (3.1) | 493 (2.2) | 107 (1.9) |
| Obstructive | 59 (12.9) | 3 (8.1) | 975 (9.6) | 3 620 (16.1) | 648 (11.2) |
| Restrictive | 162 (35.4) | 20 (54.1) | 5 686 (55.8) | 6 146 (27.3) | 2 213 (38.4) |
| Mixed | 29 (6.3) | 1 (2.7) | 1 046 (10.3) | 1 045 (4.6) | 220 (3.8) |
| **Severity** | | | | | |
| Normal | 292 (63.8) | 20 (54.1) | 4 897 (48.1) | 16 148 (71.6) | 4 226 (73.3) |
| Mild | 85 (18.6) | 9 (24.3) | 2 549 (25.0) | 3 179 (14.1) | 852 (14.8) |
| Moderate | 72 (15.7) | 7 (18.9) | 2 382 (23.4) | 2 777 (12.3) | 604 (10.5) |
| Severe | 9 (2.0) | 1 (2.7) | 363 (3.6) | 436 (1.9) | 81 (1.4) |

Values are mean (SD) for continuous variables and count (percentage) for categorical variables. Dynamic lung volume z-scores are calculated using GLI Global reference equations, while dynamic lung volume z-scores are calculated using GLI 2019 reference equations. Abbreviations: $FEV_1$ = forced expiratory volume in 1 second; FVC = forced vital capacity; GLI = Global Lung Function Initiative; TLC = total lung capacity.

**Supplementary Table S3**. Patient Characteristics in the Penn Medicine Dataset by Race and Ethnicity

| | **Asian** ($n$ = 516) | **Hispanic** ($n$ = 570) | **Non-Hispanic Black** ($n$ = 13662) | **Non-Hispanic White** ($n$ = 25759) | **Other** ($n$ = 1419) |
|---|---|---|---|---|---|
| **Age, years** | 53.0 (16.6) | 50.1 (15.2) | 53.9 (14.1) | 57.1 (14.6) | 53.1 (15.2) |
| **Sex** | | | | | |
| Male | 224 (43.4) | 231 (40.5) | 4000 (29.3) | 12284 (47.7) | 635 (44.7) |
| Female | 292 (56.6) | 339 (59.5) | 9662 (70.7) | 13475 (52.3) | 784 (55.3) |
| **Dynamic Lung Volumes, z-score** | | | | | |
| $FEV_1$ | −1.0 (1.2) | −1.2 (1.4) | −1.8 (1.1) | −1.1 (1.4) | −1.2 (1.4) |
| FVC | −0.9 (1.2) | −1.1 (1.4) | −1.7 (1.1) | −0.7 (1.3) | −1.0 (1.4) |
| $FEV_1$/FVC | −0.3 (1.3) | −0.3 (1.3) | −0.5 (1.5) | −0.7 (1.5) | −0.4 (1.4) |
| **Static Lung Volumes, z-score** | | | | | |
| TLC | −0.9 (1.3) | −1.0 (1.6) | −1.6 (1.4) | −0.6 (1.6) | −1.1 (1.6) |
| **Interpretation** | | | | | |
| Normal | 284 (55.0) | 287 (50.4) | 3883 (28.4) | 13378 (51.9) | 670 (47.2) |
| Non-Specific | 28 (5.4) | 40 (7.0) | 1345 (9.8) | 1296 (5.0) | 67 (4.7) |
| Obstructive | 65 (12.6) | 71 (12.5) | 2141 (15.7) | 5473 (21.2) | 199 (14.0) |
| Restrictive | 129 (25.0) | 160 (28.1) | 5657 (41.4) | 5128 (19.9) | 446 (31.4) |
| Mixed | 10 (1.9) | 12 (2.1) | 636 (4.7) | 484 (1.9) | 37 (2.6) |
| **Severity** | | | | | |
| Normal | 371 (71.9) | 373 (65.4) | 6219 (45.5) | 17298 (67.2) | 907 (63.9) |
| Mild | 74 (14.3) | 94 (16.5) | 3733 (27.3) | 3984 (15.5) | 268 (18.9) |
| Moderate | 70 (13.6) | 92 (16.1) | 3243 (23.7) | 3596 (14.0) | 201 (14.2) |
| Severe | 1 (0.2) | 11 (1.9) | 467 (3.4) | 881 (3.4) | 43 (3.0) |

Values are mean (SD) for continuous variables and count (percentage) for categorical variables. Dynamic lung volume z-scores are calculated using GLI Global reference equations, while dynamic lung volume z-scores are calculated using GLI 2019 reference equations. Abbreviations: $FEV_1$ = forced expiratory volume in 1 second; FVC = forced vital capacity; GLI = Global Lung Function Initiative; TLC = total lung capacity.

**Supplementary Table S4**. Patient Characteristics in the OLDW Dataset by Race and Ethnicity

| | Asian ($n = 95$) | Hispanic ($n = 187$) | Non-Hispanic Black ($n = 419$) | Non-Hispanic White ($n = 4\,046$) | Other ($n = 328$) |
|---|---|---|---|---|---|
| **Age, years** | 58.4 (16.4) | 60.8 (13.4) | 58.1 (13.1) | 63.0 (12.8) | 59.5 (14.7) |
| **Sex** | | | | | |
| Male | 34 (35.8) | 66 (35.3) | 146 (34.8) | 1 710 (43.3) | 153 (46.6) |
| Female | 61 (64.2) | 121 (64.7) | 273 (65.2) | 2 336 (57.7) | 175 (53.4) |
| **Dynamic Lung Volumes, z-score** | | | | | |
| $FEV_1$ | −0.4 (0.9) | −0.6 (1.3) | −1.4 (1.2) | −0.6 (1.4) | −0.5 (1.3) |
| FVC | −0.2 (0.8) | −0.3 (1.1) | −1.4 (1.1) | −0.2 (1.3) | −0.1 (1.2) |
| $FEV_1$/FVC | −0.5 (0.9) | −0.4 (1.5) | −0.3 (1.2) | −0.7 (1.6) | −0.7 (1.2) |
| **Static Lung Volumes, z-score** | | | | | |
| TLC | −0.7 (0.9) | −0.8 (1.3) | −1.9 (1.3) | −0.4 (1.4) | −0.4 (1.3) |
| **Interpretation** | | | | | |
| Normal | 73 (76.8) | 117 (62.6) | 162 (38.7) | 2 679 (66.2) | 220 (67.1) |
| Non-Specific | $< 11$ | $< 11$ | $< 11$ | 17 (0.4) | $< 11$ |
| Obstructive | $< 11$ | 25 (3.4) | $> 11$ | 703 (17.4) | 56 (17.1) |
| Restrictive | 14 (14.7) | 40 (21.4) | 198 (47.3) | 564 (13.9) | 45 (13.7) |
| Mixed | $< 11$ | $< 11$ | 31 (7.4) | 83 (2.1) | $< 11$ |
| **Severity** | | | | | |
| Normal | 84 (88.4) | 150 (80.2) | 255 (60.9) | 3 111 (76.9) | 259 (79.0) |
| Mild | $< 11$ | 21 (11.2) | 81 (19.3) | 479 (11.8) | 43 (13.1) |
| Moderate | $< 11$ | $> 11$ | $> 11$ | 410 ( 10.1) | $> 11$ |
| Severe | $< 11$ | $< 11$ | $< 11$ | 46 (1.1) | $< 11$ |

Values are mean (SD) for continuous variables and count (percentage) for categorical variables. Dynamic lung volume z-scores are calculated using GLI Global reference equations, while dynamic lung volume z-scores are calculated using GLI 2019 reference equations. To protect patient privacy, cells containing 10 or fewer patients are listed as <11. Analogous adjustments are made to other cells to prevent the number of patients from being calculated from the column sum. Abbreviations: $FEV_1$ = forced expiratory volume in 1 second; FVC = forced vital capacity; GLI = Global Lung Function Initiative; OLDW = Optum Labs Data Warehouse; SD = standard deviation; TLC = total lung capacity.

**Supplementary Table S5**. Patient Characteristics by Pulmonary Diagnostic Lab

| | **Penn Medicine** | | |
|---|---|---|---|
| | **Lab 1** ($n = 28\,503$) | **Lab 2** ($n = 6\,739$) | **Lab 3** ($n = 6\,684$) |
| **Age, years** | 54.9 (14.6) | 57.2 (14.1) | 58.2 (14.5) |
| **Sex** | | | |
| Male | 12 314 (43.2) | 2 487 (36.9) | 2 573 (38.5) |
| Female | 16 189 (56.8) | 4 252 (63.1) | 4 111 (61.5) |
| **Race and Ethnicity** | | | |
| Asian | 288 (1.0) | 106 (1.6) | 122 (1.8) |
| Hispanic | 286 (1.0) | 76 (1.0) | 208 (2.9) |
| Non-Hispanic Black | 7 828 (27.5) | 3 742 (55.5) | 2 092 (31.3) |
| Non-Hispanic White | 18 811 (66.0) | 2 765 (41.0) | 4 183 (62.6) |
| Other | 1290 (4.5) | 50 (0.7) | 79 (1.2) |
| **Dynamic Lung Volumes, z-score** | | | |
| $FEV_1$ | −1.4 (1.9) | −1.5 (1.8) | −1.2 (1.8) |
| FVC | −1.0 (1.8) | −1.3 (1.7) | −0.9 (1.3) |
| $FEV_1$/FVC | −0.7 (1.5) | −0.5 (1.5) | −0.6 (1.5) |
| **Static Lung Volumes, z-score** | | | |
| TLC | −1.0 (1.6) | −1.3 (1.4) | −0.3 (1.5) |
| **Interpretation** | | | |
| Normal | 12 619 (44.3) | 2 470 (36.7) | 3 413 (51.1) |
| Non-Specific | 1 544 (5.4) | 435 (6.5) | 797 (11.9) |
| Obstructive | 5 432 (19.1) | 1 158 (17.2) | 1 359 (20.3) |
| Restrictive | 8 082 (28.4) | 2 400 (35.6) | 1 038 (15.5) |
| Mixed | 826 (2.9) | 276 (4.1) | 77 (1.2) |
| **Severity** | | | |
| Normal | 17 269 (60.6) | 3 678 (54.6) | 4 221 (63.2) |
| Mild | 5 347 (18.8) | 1 496 (22.2) | 1 310 (19.6) |
| Moderate | 4 762 (16.7) | 1 392 (20.7) | 1 048 (15.7) |
| Severe | 1 125 (3.9) | 173 (2.6) | 105 (1.6) |

Values are mean (SD) for continuous variables and count (percentage) for categorical variables. Dynamic lung volume z-scores are calculated using GLI Global reference equations, while dynamic lung volume z-scores are calculated using GLI 2019 reference equations. Abbreviations: $FEV_1$ = forced expiratory volume in 1 second; FVC = forced vital capacity; GLI = Global Lung Function Initiative; SD = standard deviation; TLC = total lung capacity.

**Supplementary Table S6**. Test Characteristics by Dataset

| Dataset | Sensitivity (%) | Specificity (%) | NPV (%) |
|---|---|---|---|
| Johns Hopkins Medicine | 53.7 (53.0 to 54.5) | 92.1 (91.8 to 92.5) | 72.9 (72.4 to 73.5) |
| Penn Medicine | 70.2 (69.4 to 71.0) | 84.2 (83.7 to 84.6) | 86.7 (86.3 to 87.1) |
| OLDW | 52.5 (49.3 to 55.6) | 93.2 (92.4 to 94.0) | 89.0 (88.1 to 90.0) |

NPV = negative predictive value; OLDW = Optum Labs Data Warehouse.

**Supplementary Table S7**. Test Characteristics by Pulmonary Diagnostic Lab

| Dataset | Lab | Sensitivity (%) | Specificity (%) | NPV (%) |
|---|---|---|---|---|
| Penn Medicine | Lab 1 | 69.2 (68.3 to 70.2) | 86.1 (85.6 to 86.6) | 86.0 (85.5 to 86.5) |
| | Lab 2 | 70.0 (68.2 to 71.7) | 79.2 (77.9 to 80.5) | 80.0 (78.8 to 81.3) |
| | Lab 3 | 78.7 (76.2 to 81.1) | 80.9 (79.8 to 81.9) | 95.0 (94.3 to 95.6) |

Abbreviations: NPV = negative predictive value.

**Supplementary Table S8**. Test Characteristics by Race and Ethnicity

| **Dataset** | **Race and Ethnicity** | **Sensitivity (%)** | **Specificity (%)** | **NPV (%)** |
|---|---|---|---|---|
| | Asian | 64.4 (57.2 to 71.2) | 92.9 (89.1 to 95.7) | 78.5 (73.5 to 82.9) |
| | Hispanic | 57.1 (34.0 to 78.2) | 87.5 (61.7 to 98.4) | 60.9 (38.5 to 80.3) |
| Johns Hopkins Medicine | Non-Hispanic Black | 60.5 (59.3 to 61.6) | 83.8 (82.6 to 85.1) | 52.1 (50.8 to 53.5) |
| | Non-Hispanic White | 49.7 (48.5 to 50.8) | 93.3 (92.9 to 93.7) | 79.8 (79.2 to 80.4) |
| | Other | 46.2 (44.2 to 48.2) | 95.5 (94.7 to 96.2) | 70.8 (69.5 to 72.2) |
| | Asian | 69.8 (61.4 to 77.3) | 87.5 (83.8 to 90.7) | 88.7 (85.0 to 91.7) |
| | Hispanic | 72.1 (64.8 to 78.7) | 87.2 (83.5 to 90.3) | 87.8 (84.2 to 90.9) |
| Penn Medicine | Non-Hispanic Black | 73.4 (72.2 to 74.5) | 71.1 (70.1 to 72.2) | 75.8 (74.8 to 76.8) |
| | Non-Hispanic White | 66.9 (65.6 to 68.1) | 88.6 (88.2 to 89.0) | 90.6 (90.2 to 91.0) |
| | Other | 67.5 (63.1 to 71.7) | 88.2 (86.0 to 90.2) | 84.0 (81.6 to 86.3) |
| | Asian | 25.0 (7.3 to 52.4) | 100.0 (95.4 to 100.0) | 86.8 (78.1 to 93.0) |
| | Hispanic | 36.4 (22.4 to 52.2) | 95.1 (90.2 to 98.0) | 82.9 (76.3 to 88.3) |
| OLDW | Non-Hispanic Black | 55.5 (48.8 to 62.0) | 88.4 (83.0 to 92.6) | 62.2 (56.1 to 68.0) |
| | Non-Hispanic White | 52.9 (48.9 to 56.8) | 93.1 (92.2 to 93.9) | 91.2 (90.2 to 92.1) |
| | Other | 56.9 (42.2 to 70.7) | 95.3 (92.1 to 97.5) | 92.3 (88.6 to 95.1) |
| | Asian | 64.7 (59.5 to 69.8) | 90.9 (88.5 to 92.9) | 84.3 (81.6 to 86.8) |
| | Hispanic | 64.1 (57.7 to 70.2) | 89.2 (86.4 to 91.7) | 85.4 (82.3 to 88.2) |
| All | Non-Hispanic Black | 66.5 (65.7 to 67.3) | 75.4 (74.6 to 76.2) | 65.2 (64.4 to 66.0) |
| | Non-Hispanic White | 57.0 (56.2 to 57.8) | 90.8 (90.6 to 91.1) | 85.9 (85.6 to 86.3) |
| | Other | 49.8 (48.0 to 51.6) | 94.0 (93.3 to 94.7) | 74.1 (73.0 to 75.3) |

Abbreviations: NPV = negative predictive value; OLDW = Optum Labs Data Warehouse.

**Supplementary Table S9**. NPV in Non-Hispanic White Patients with Different Reference Equations

| **Reference Equations for Dynamic Lung Volumes** | **Reference Equations for Static Lung Volumes** | | |
|---|---|---|---|
| | Crapo et al.[1] | ECSC[2] | GLI 2019[3] |
| Knudson et al. and Enright et al.[4,5] | 92.2 (92.0 to 92.5) | 86.0 (85.6 to 86.3) | 86.7 (86.4 to 87.1) |
| Hankinson et al.[6] | 95.8 (95.5 to 96.0) | 91.7 (91.3 to 92.0) | 92.7 (92.4 to 93.0) |
| GLI 2012[7] | 93.1 (92.8 to 93.3) | 87.8 (87.4 to 88.1) | 89.0 (88.6 to 89.3) |
| GLI Global[8] | 90.7 (90.4 to 91.0) | 84.7 (84.3 to 85.1) | 85.4 (85.1 to 85.8) |

Abbreviations: ECSC = European Coal and Steel Community; GLI = Global Lung Function Initiative.

**Supplementary Table S10**. Test Characteristics with Race-Specific and Race-Neutral Reference Equations

| Race and Ethnicity | Reference Equations | Sensitivity (%) | Specificity (%) | NPV (%) |
|---|---|---|---|---|
| All | Race Specific | 59.5 (58.9 to 60.0) | 85.2 (84.9 to 85.5) | 78.8 (78.4 to 79.1) |
| | Race Neutral | 60.9 (60.3 to 61.4) | 87.6 (87.3 to 87.9) | 79.8 (79.5 to 80.1) |
| Asian | Race Specific | 71.2 (66.0 to 76.0) | 87.1 (84.3 to 89.6) | 85.5 (82.6 to 88.1) |
| | Race Neutral | 66.7 (61.3 to 71.7) | 88.1 (84.4 to 91.2) | 89.3 (85.7 to 92.3) |
| Hispanic | Race Specific | 75.2 (67.3 to 82.0) | 89.8 (87.1 to 92.0) | 87.3 (83.6 to 90.5) |
| | Race Neutral | 70.5 (63.5 to 76.8) | 87.2 (83.6 to 90.3) | 86.4 (82.7 to 89.5) |
| Non-Hispanic Black | Race Specific | 48.6 (47.7 to 49.4) | 85.7 (85.0 to 86.3) | 58.1 (57.3 to 58.8) |
| | Race Neutral | 66.7 (65.9 to 67.5) | 75.2 (74.4 to 76.0) | 65.3 (64.4 to 66.1) |
| Non-Hispanic White | Race Specific | 71.1 (70.4 to 71.9) | 84.1 (83.7 to 84.5) | 89.0 (88.6 to 89.3) |
| | Race Neutral | 57.2 (56.4 to 58.1) | 90.6 (90.3 to 90.9) | 85.4 (85.1 to 85.8) |
| Other | Race Specific | 54.7 (52.9 to 56.6) | 93.1 (92.3 to 93.8) | 75.1 (73.9 to 76.2) |
| | Race Neutral | 49.7 (47.9 to 51.5) | 93.9 (93.1 to 94.6) | 73.2 (72.0 to 74.4) |

Race-specific reference equations use GLI 2012 for dynamic lung volumes and GLI 2019 for static lung volumes. Race-neutral reference equations use GLI Global for dynamic lung volumes and GLI 2019 for static lung volumes. Abbreviations: GLI = Global Lung Function Initiative; NPV = negative predictive value; PPV = positive predictive value.

**Supplementary Figure S1.** Flow Diagram for the Johns Hopkins Medicine Dataset

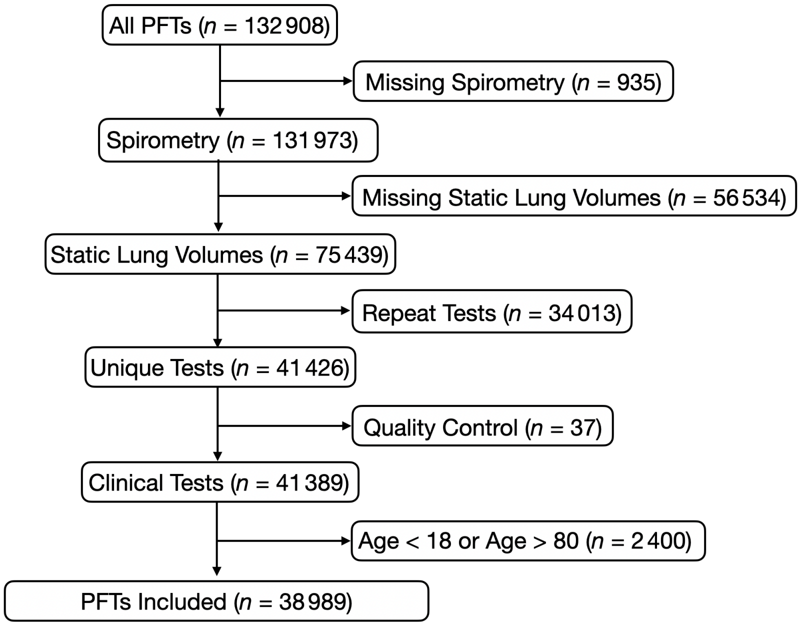

**Supplementary Figure S2.** Flow Diagram for the Penn Medicine Dataset

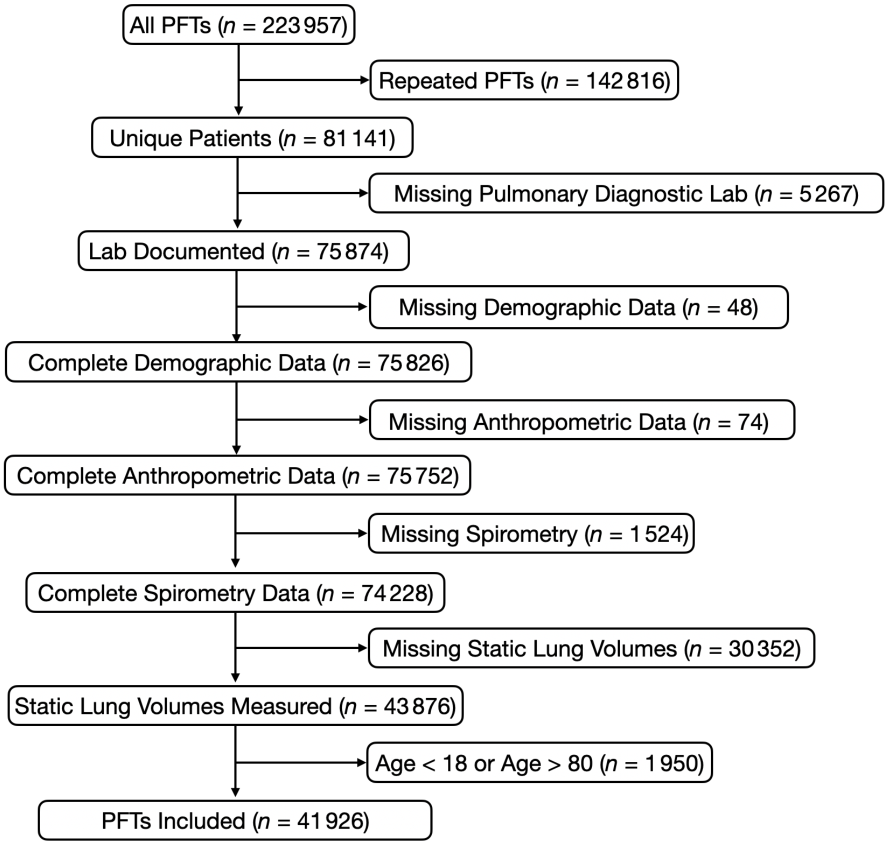

**Supplementary Figure S3.** Flow Diagram for the OLDW Dataset

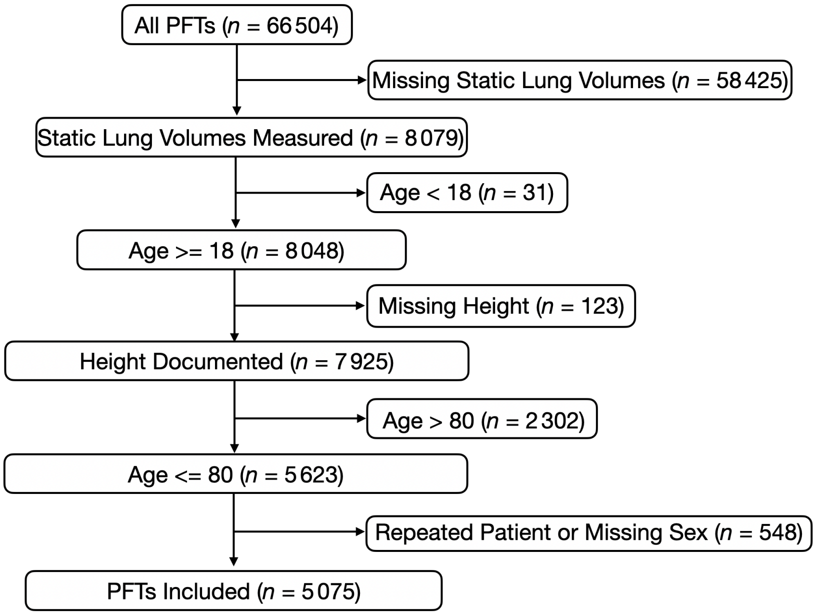
